# On Machine Learning-Based Short-Term Adjustment of Epidemiological Projections of COVID-19 in US

**DOI:** 10.1101/2020.09.11.20180521

**Authors:** Sarah Kefayati, Fred Roberts, Sayali Pethe, Xuan Liu, Hu Huang, Vishrawas Gopalakrishnan, Piyush Madan, Jianying Hu, Prithwish Chakraborty, Raman Srinivasan, Ajay Deshpande, Gretchen Jackson

## Abstract

Epidemiological models have provided valuable information for the outlook of COVID-19 pandemic and relative impact of different mitigation scenarios. However, more accurate forecasts are often needed at near term for planning and staffing. We present our early results from a systemic analysis of short-term adjustment of epidemiological modeling of COVID 19 pandemic in US during March-April 2020. Our analysis includes the importance of various types of features for short term adjustment of the predictions. In addition, we explore the potential of data augmentation to address the data limitation for an emerging pandemic. Following published literature, we employ data augmentation via clustering of regions and evaluate a number of clustering strategies to identify early patterns from the data.

From our early analysis, we used CovidActNow as our underlying epidemiological model and found that the most impactful features for the one-day prediction horizon are population density, workers in commuting flow, number of deaths in the day prior to prediction date, and the autoregressive features of new COVID-19 cases from three previous dates of the prediction. Interestingly, we also found that counties clustered with New York County resulted in best preforming model with maximum of *R^2^=* 0.90 and minimum of *R*^2^=0.85 for state-based and COVID-based clustering strategy, respectively.

## INTRODUCTION

The novel Coronavirus, SARS-CoV-2, was first detected in Wuhan, China on December 31, 2019 and by early January 2020, it spread to 21 countries. The first case in the United States (US) was reported on January 21, 2020 in Snohomish County in the state of Washington [1]. By the middle of March 2020, the number of COVID-19 infected cases started to peak up in the US [2], inducing panic about potential shortages of hospital capacities and supplies.

As more cases were detected in the US, many epidemiological models, which had been mainly developed for other pandemics such as influenza and season-long forecasting, started to adjust to and project COVID-19 growth patterns. Due to the critical role of forecasting COVID-19 in helping policymaker for future planning, the Center of Disease Control and Prevention (CDC) has initiated an effort for forecasting in form of challenge with the participation of several of the epidemiological models [3].

Although overall epidemiological models have been useful for understanding the future outlook of COVID-19 pandemic, they often have been criticized for overestimating the projections and inducing uncertainties [4]. The limitations in predicting future cases of COVID-19 stems from a combination of limited understanding and a lack of data about its infectious spread pattern due to its novel nature. Beside data limitations, region-specific factors were often not accounted-for in many published epidemiological models of the COVID-19 pandemic. Factors such as age distribution, comorbidities, and pre-excising conditions, as well as socioeconomic factors, are expected to play role in the severity and duration of this disease.

Given the importance of COVID-19 forecasting and considering the majority of epidemiological models are best suited for long-term projections, in this work rather than developing a new forecasting model, we aim at improving the overestimated projected numbers from epidemiological models on a short-term basis based on machine learning. In particular, we conducted a systematic analysis to study the importance of various data elements for our short-term prediction. Our predictive modeling included a carefully selected 40 geo-specific features for the US counties.

A previous study by Liu et al. on machine learning-based predictions of COVID-19 outbreak in China, showed that modeling for the regions clustered together based on geo-specific similarities resulted in improved predictive performance for majority of China provinces [5]. In this work, we studied four different clustering strategies, primarily based on demographic similarities, COVID activity trends, state boundaries, as well as a national cluster, to examine the effects of these boundary conditions in predictive power of our model.

Our main contribution is investigating the role of machine learning to adjust the short-term epidemiological projections with the ultimate aim of helping counties and hospitals better plan their resources. We are also investigating which additional region-specific features play a role in short-term adjustments of the COVID-19 projections.

## 1 RESULTS

For historical epidemiological projection, we used the CovidActNow model as one of the early epidemiological models that became opensource [6]. For the data augmentation purpose, we considered four clustering strategies: one national cluster including all the counties in our training set, second clustering based on counties state boundary, and another two clustering strategies were based on counties similarities in COVID-19 spread characteristics and demographic information as described in the Methods section.

The epidemiological historical projections were one-day ahead on any given day during March 23^rd^-April 20^th^, 2020 period. We trained an individual date-dependent models for each of the clusters within each clustering strategy. The out-of-sample test sets were evaluated from April 6^th^ to April 19^th^, 2020 individually each with the date-appropriate trained model (the total of 14 date-dependent models for each cluster). The comparison and the impact of the four different clustering boundary conditions is shown in Table 1.

**Table 1:**
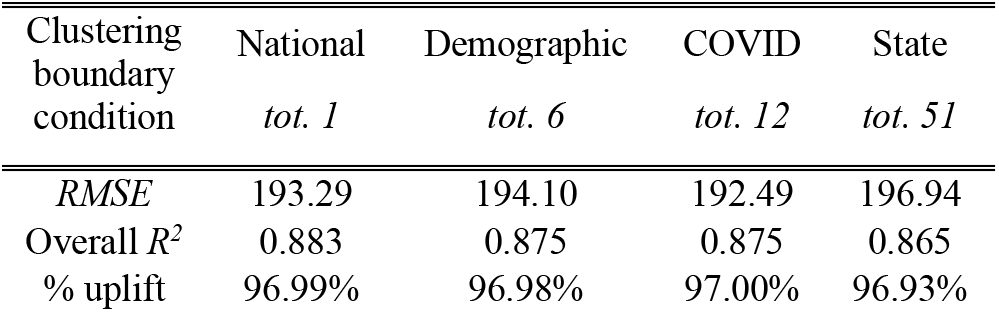
Comparison of four clustering strategies. *RMSE* values are derived by comparing our model predictions to the ActNow model ground truth values. The uplift percentage is derived based on ActNow projections *(RMSE =* 6427.27)

Regardless of the clustering strategy, our one-day-ahead prediction showed ~97% uplift compared to the epidemic projection with the same one-day-ahead horizon. Overall, the performance of the models based on each clustering strategy was similar.

Also consistent across all four different clustering strategies, the best model performance was achieved for the cluster in which New York County was included. Figure 1 shows the comparison of our model output to the CovidActNow projected “all infected” cases both compared with the ground truth (i.e., estimated all infected) for three example counties which clustered together with New York County for two different clustering strategies. Both COVID-based and demographic-based clustering achieved 97% uplift compared to the epidemiological projections for the cluster model that included New York County.

**Figure 1:**
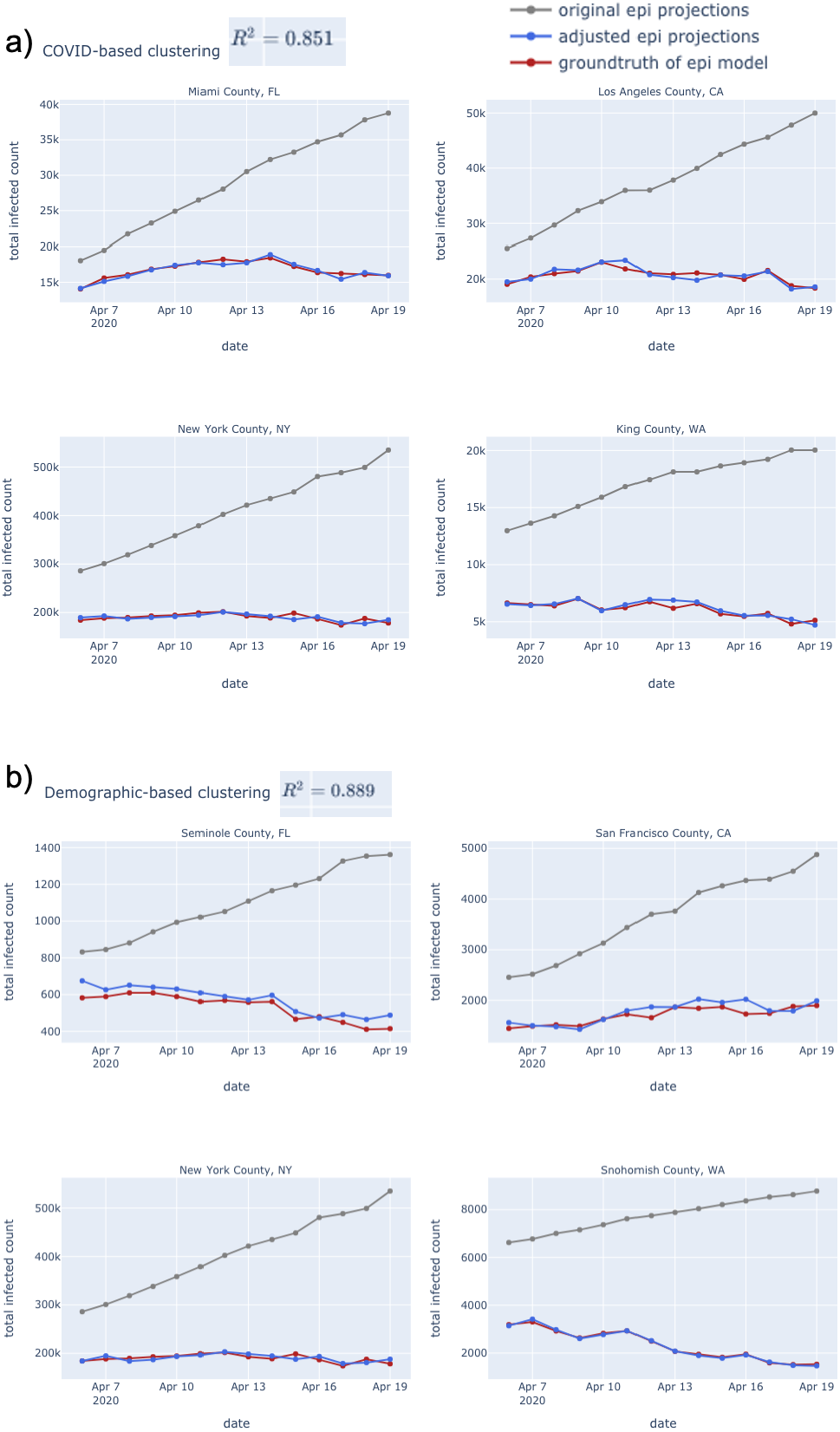
One-day-horizon predictions from our machine learning model and from the CovidActNow epidemiological model both compared to the ground truth for two different clustering strategies: a) one of the 12 clusters of the COVID-based clustering with 49 counties in the cluster, and b) one of the 6 clusters of the demographic-based clustering with 259 counties in the cluster.

Closer assessment of two major counties of New York and Los Angeles further revealed the similarities of four different clustering strategies, particularly for New York County as shown in Figure 2. The best performing cluster model for New York County was found to be state-based clustering with *R^2^= 0.90*. Los Angeles County was obtained from the COVID-based clustering method, which follows similar trajectory as the national clustering with comparable *RMSE* (676.34 and 695.61, respectively). State-clustered model showed highest deviation with *RMSE* = 1249.78, followed by second highest *RMSE* = 723.75 from the demographic-based clustering model. We found that the level of COVID-19 inactivity or low activity in the cluster impacted the performance for the corresponding model. For example, comparing two of the clusters containing Los Angeles County from two different clustering strategies, the state-based cluster contained 36% zero cases as opposed to only 2% zero cases in the COVID-based cluster.

**Figure 2:**
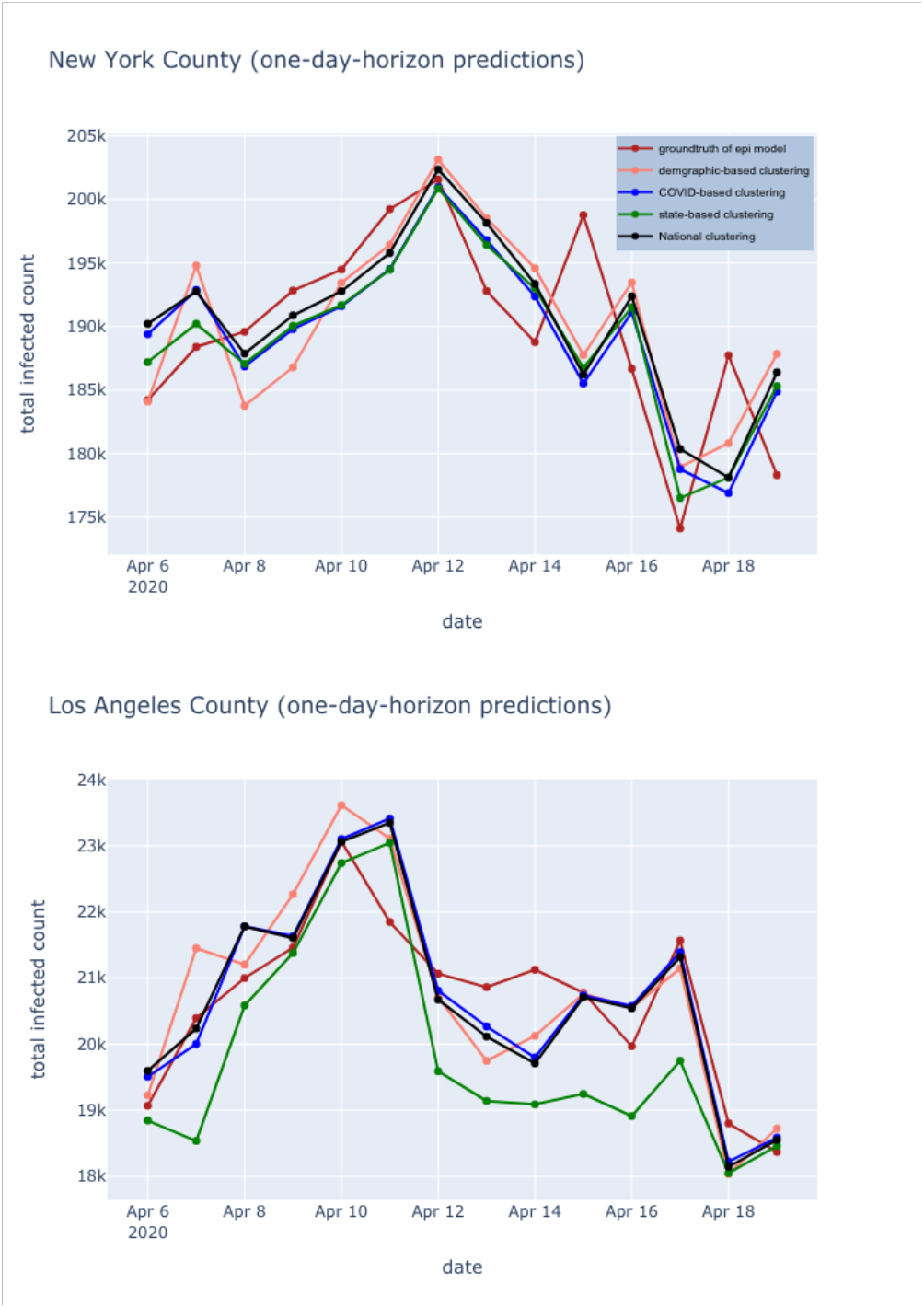
One-day-horizon predictions of our models trained individually according to the clustering strategy for two of the major counties.

As LASSO gives spare weights by driving small weights to zero, we can detect the most predictive features of our model. As walk-forward split expands by every date and more training data gets included in the split, fewer features become prominent. For the national model, as shown in Figure 3, the most impactful features for the last trained model were population density, and the autoregressive features of new COVID cases from three previous date of the prediction.

**Figure 3:**
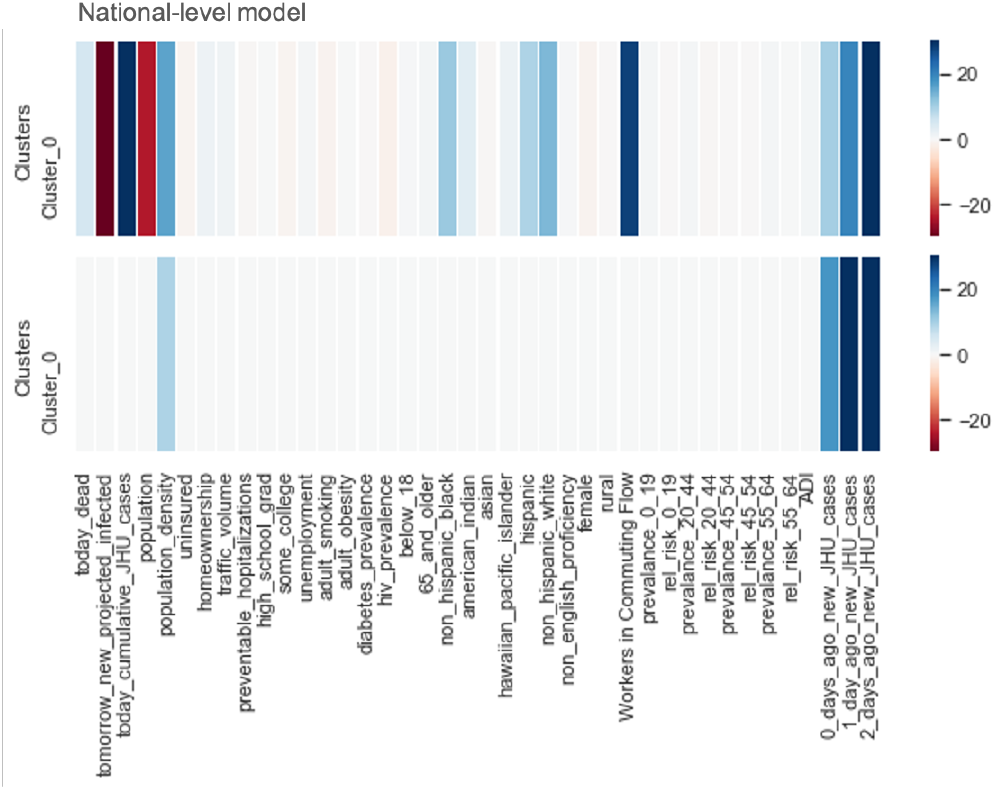
LASSO selected features for the first (top panel) and last (bottom panel) trained models (out of 14 date-dependents models) trained for all of the counties included in our model (total of 1578 counties). Although majority are small values, none of the features in the first trained model have zero coefficients. The last model has 36 features with coefficients value of zero displayed for better comparison.

In the first trained model (with the least training data), in addition to population density and autoregressive features, workers in commuting flow and cumulative number of cases had positive predictive effects on predicting the next day’s infected cases. In contrast, population and next day projected cases from the CovidActNow output had negative predictive power. We speculate the negative predictive power of CovidActNow is due to its increasing trend as opposed to ground truth that has downward trend generally. It is also important to note for the first trained model (first expanding window iteration) with limited training data, LASSO attempts to distribute the weights to more features. As the training data expands in the last trained window, the effect of CovidActNow projections is eliminated. Other features had small weights (either negative or positive) and eventually weighted to zero for the last trained model. Similarly, we found that population density and three autoregressive features were the most important (non-zero and positive) features for the majority including best performing clusters, for other clustering strategies. For the demographic-based clustering due to homogeneous distribution of the, regardless of the date of trained model, none of the demographic features carried any weight with the exception of the population density and workers in commuting flow. Overall, no particular pattern was detected for the role of race and sex with few exceptions. For instance, the proportion of non-Hispanic black population showed positive predictive impact for the State of Wisconsin with the state-based clustering approach. For feature importance map of other three clustering strategy please refer to APPENDIX II.

## 2 Methods

For the purpose of data augmentation, we considered grouping counties four different ways to ensure the that the scarcity of available training data during early period of COVID-19 pandemic (window considered March 23^rd^-April 20^th^, 2020) did not impact the model performance. One grouping included counties of each of the US states, yielding 51 individual trained models. Another national approach involved grouping all the counties together and training one model for all. The two other groupings were conducted by clustering the counties based on their similarities in COVID-19 spread characteristics and demographic information as described below.

### 2.1 Clustering Strategy

Clustering was conducted based on agglomerative hierarchal clustering algorithm (scikit-learn package), which is a bottom-up approach merging counties based on their similarities until reaching one big cluster. The optimal number of clusters was obtained by maximizing Calinski-Harabasz score.

The counties were clustered based on their demographic information for which the optimization yielded 6 clusters. Features included race, ethnicity, gender, elderly (age > 65 years) and young (age < 18 years) population, total and density of population, county traffic volume, county average commute flow, as well as Area Deprivation Index, rankings of counties by socioeconomic status disadvantage [7].

Counties were also separately clustered based on time-varying COVID-19 characteristics, resulting in 12 dissimilar clusters. Features included reciprocal doubling time based on the growth during the selected time window, the pattern of the growth curve (logistic, exponential or none), cumulative cases on April 20^th^, 2020, and number of days stayed at home since the stay-at-home order for each county. Maps of the clustered US counties based on their demographic and COVID-19 activity similarities are available in APPENDIX I.

### 2.2 Epidemiological Projections

The Coronavirus Act Now model is SEIR model, one of the main groups of epidemiological models used by epidemiologists and researchers to project the evolution of a disease. The model works by categorizing the population at various states and modeling as the population moves through susceptible (S), individuals becoming exposed (E) to the virus, then infected (I), and infected population either recovering (R) or dying (D). To model hospitalization and need for ICU bed, the infected cases were categorized into three levels of disease severity: mild (no hospitalization), moderate (requiring hospitalization), or severe (requiring ICU bed)^1^.

For the earlier version of the ActNow model, caseload data (number of confirmed cases and death) were updated daily from the Johns Hopkins University (JHU) Center for Systems Science and Engineering’s Coronavirus Tracking Dashboard [8] with the county-level data becoming available as of March 22^nd^, 2020.

We used “all infected” projection (including mild, moderate, and severe cases) output of the CovidActNow model to compare with our machine learning forecasting. In order to compare our model results with CovidActNow, we used the same ground truth as CovidActNow [6]. In the earlier version of the ActNow model^1^, the ground truth for the “all infected” (i.e. estimated all infected) was derived as follow: “estimated recovered” was estimated from actual COVID cases report (JHU) shifted by 13 days. The “estimated recovered” cases together with total number of deceased populations on a given date gives an estimate of active cases. Due to lack of reported data on hospitalization at the county level, CovidActNow then estimates that a quarter of the “estimated active” cases are hospitalized. And finally, that “estimated hospitalization” is about 7.3% of “estimated all infected” (mild, moderate, and severe) cases. Thus, we used “estimated all infected” as the ground truth to compare our forecasting to the ActNow model projections.

The one-day-horizon CovidActNow historical projections were obtained by running a model for every day during March 23^rd^-April 20^th^, 2020 period and selecting the next-day projections yielding 28 days datapoints. Since the ActNow model produced several intervention scenarios, for each date, we selected the output that matched the actual in-place intervention. We obtained the intervention policies for each state from The New York Times website. For some of the states, the policies were not held state-wide, thus scraped those individual county policies from various news outlets.

### 2.3 Predictive Model

For the predictive modeling, we fitted our data to a LASSO model, a multivariate linear model with L1-norm regularization (penalizing the absolute sum of the model coefficients). The feature vector included both time-dependent and static features for each county. The time-dependent were COVID-19 dependent features both from the epidemiological model output and from JHU reported parameters. The time-independent features were demographic and characteristics of each county. In total, static and time-varying, 40 features were included in our model. For the list of the features and their sources please refer to APPENDIX III.

The number of new confirmed COVID-19 cases for the next day was then predicted based on:

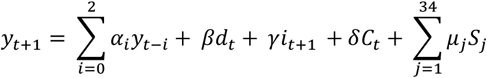

Where *y_t+_*_1_, *y_t_, y_t−_*_1_, and *y_t_*_-2_ are the new confirmed cases for the next day, same date, the day before, and two days prior of date t. *d_t_* refers to the new death number at date t; *i_t+_*_1_ is the new “all infected” cases projected for next day at date t from the epidemiological model, and *C_t_* is the cumulative number of COVID-19 cases on date t. *S_j_* refers to collection of 34 static features including, population density, county commute flow, age, gender, race, socioeconomic features such as unemployment and Area Deprivation Index, as well as disease prevalence and comorbidities.

Due to time-series nature and daily update of the data, a walk-forward with expanding window validation fashion was considered resulting in 15 splits with one day horizon and initial training window of 10 days. Since the split was performed according to the date of data, each cluster yielded 15 sets consistently; however, depending on number of counties in each cluster, different train-test splits were expected for each cluster. The total of 1578 counties were included in this study each with feature vector of size 40. To compare the predicted outcome of our model with the epidemiological projected cases, the predicted new confirmed cases were then summed with the JHU-reported “cumulative number of cases” from the previous date, and then converted to “estimated all infected” according to the conversion used in the CovidActNow model.

The best model parameters, including LASSO alpha, were selected in a Grid search manner. To simulate the real-life scenario in which the last trained model needs to be updated daily as the new data comes in, the model trained on the first n-1 splits (n=2, 3, 4, … 15), was evaluated on the test set of nth split (i.e. out-of-sample test set).

## 3 CONCLUSIONS

Our machine learning results showed significant improvement over the epidemiological projection with a one-day prediction horizon. Although small, the changes in model performance was detectable based on different clustering strategies. Assessing New York County, for example, we found state-based clustering achieved the highest performance for the state of New York *(R^2^* = 0.90). However, state-based clustering resulted in lowest performance for overall counties compared to other strategies (Table 1). This finding suggests an advantage to considering training individual models based on the geographic region.

The results shown for our model are from March-April 2020 period of time, a time of dramatic increases in COVID-19 spread in major cities with subsequent implementation of mitigation strategies, such as stay-at-home policies with California first to enforce such a policy on March 19^th^, 2020. Our findings of different clustering strategies and feature importance were likely due to the highly dynamic nature of infectious spread and local policies at this time. Moreover, as more data has become available over time, epidemiological models are also improving and more scenarios are considered in the modeling methods, such as the impact of asymptomatic cases in the recent version of the ActNow model.

It is also important to note that for this work, our prediction is short and limited to one-day horizon. Although, it is not expected that all of the 40 features that we have included have predictive power, we suspect with longer prediction horizon, demographic and geo-specific features will have important roles.

With initially flattening and now new rises in the pandemic curve, we plan to expand our modeling prediction horizon to as long as two weeks ahead. Some of the challenges that we foresee with longer prediction horizon is the policy changes as the states would go through different phases of reopening. We suspect for longer prediction horizon, more COVID-specific trends for each state as well as information about the upcoming policies can improve the predictive powers. Also, we intend to incorporate several epidemiological models’ outputs to study their difference and impact in our modeling.

## Data Availability

The data used in model building are all publicly available data except comorbidity data that is IBM proprietary.

## APPENDIX I: Clustering of US Counties

**Supplementary Figure 1:**
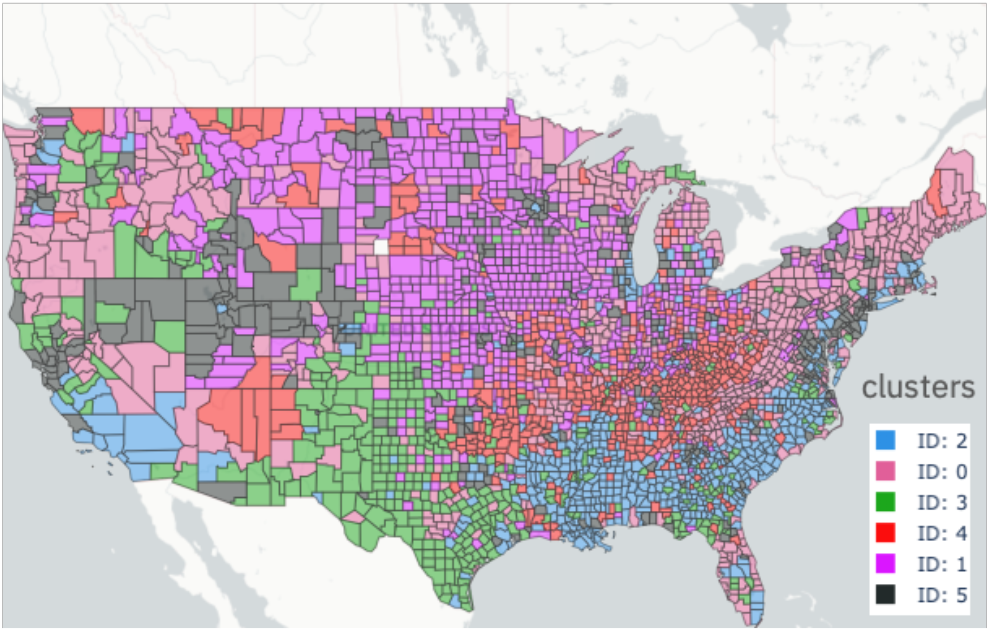
US counties clustering based on their demographic similarities. For the reference, New York county is in cluster 5.

**Supplementary Figure 2:**
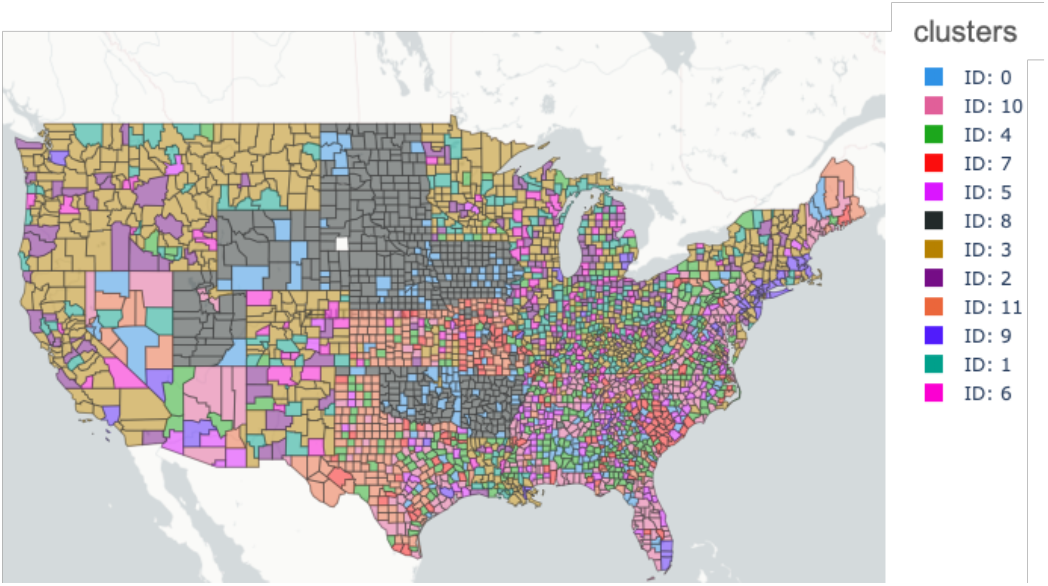
US counties clustering based on their COVID-19 activity similarities during March 23^rd^-April 20^th^ 2020. For the reference, New York county belongs to cluster 9.

## Supplementary APPENDIX II: LASSO Selected Features

**Supplementary Figure 3:**
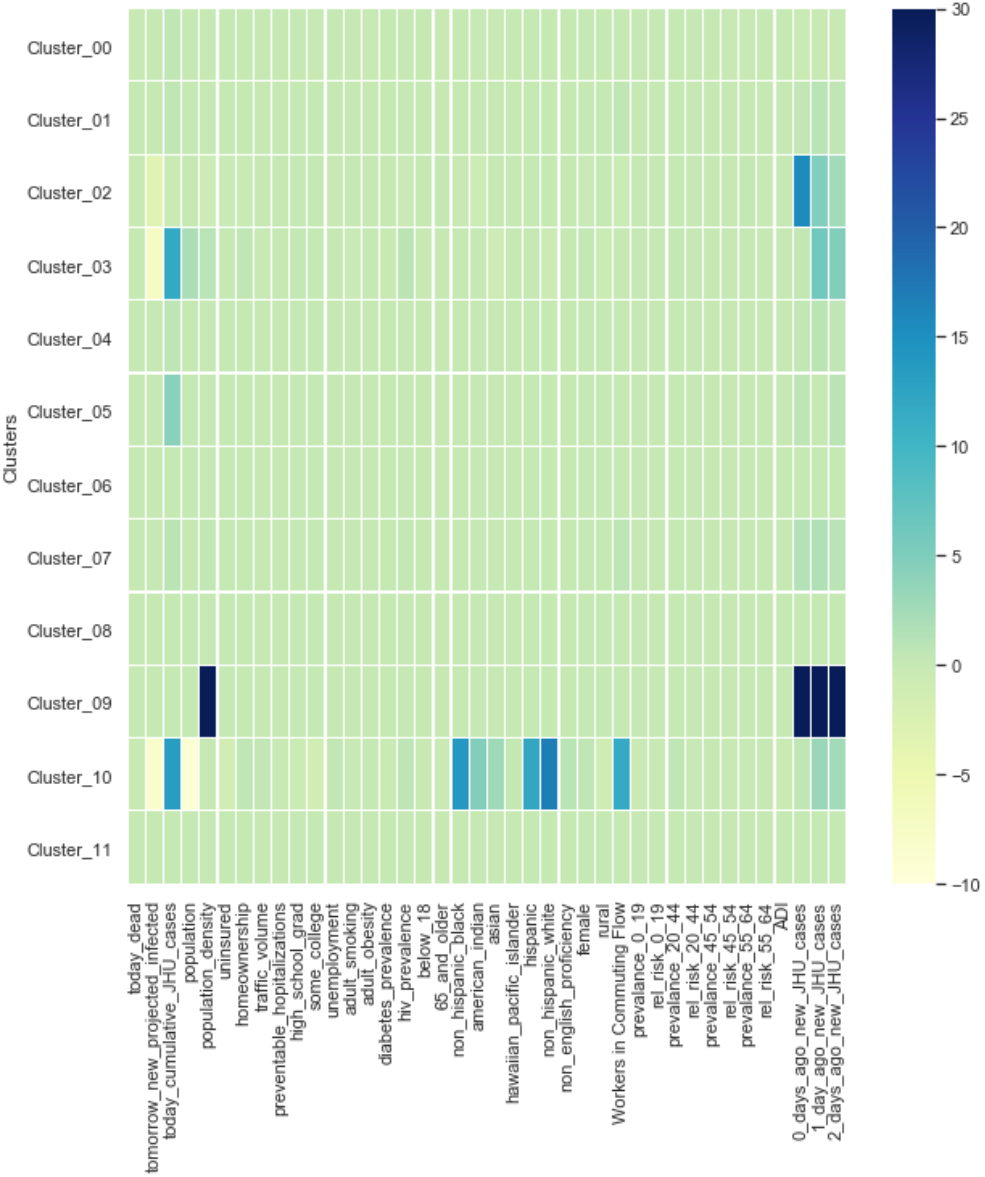
LASSO selected features for the last trained model (out of 14 date-dependents models) of each cluster trained for the counties clustered together based on similarities of COVID characteristics. New York County is part of cluster 9, yielding best preforming model.

**Supplementary Figure 4:**
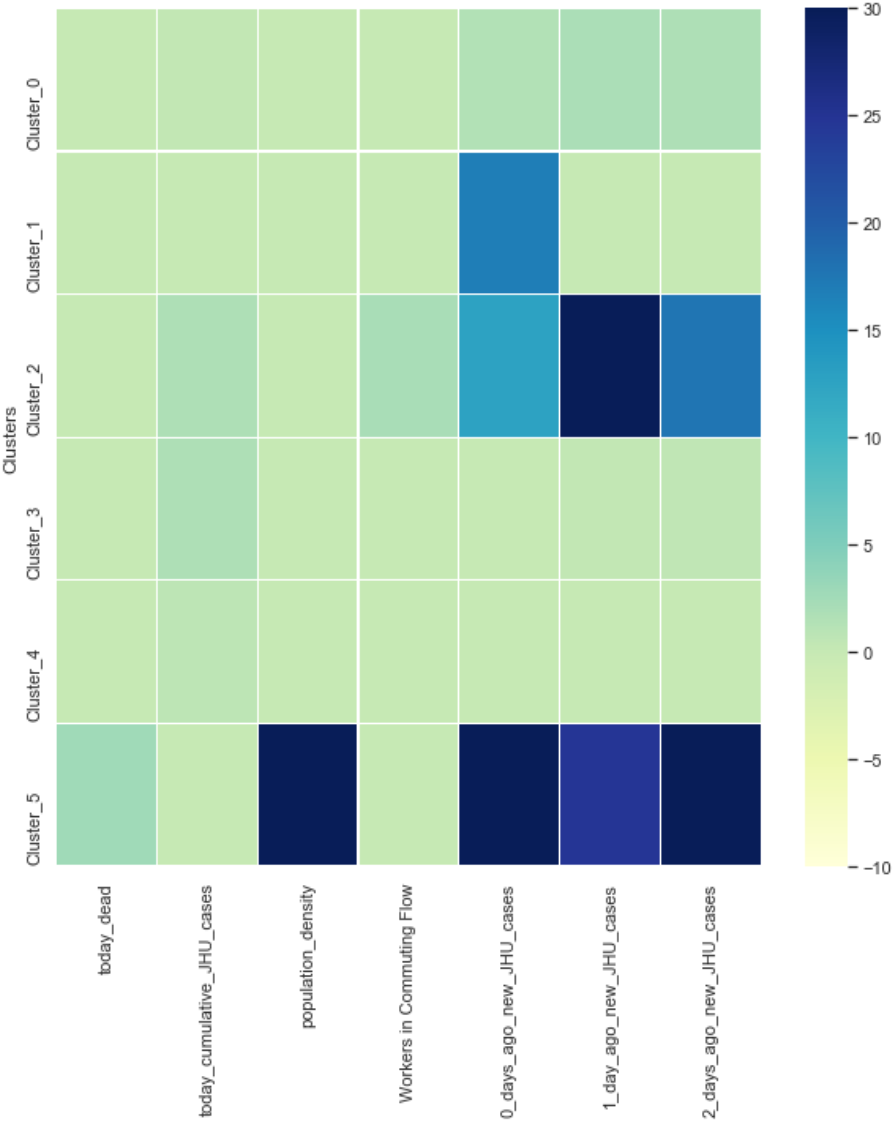
LASSO selected features for the last trained model (out of 14 date-dependents models) of each cluster trained for the counties clustered together based on their demographic similarities. New York County is part of cluster 5, yielding best preforming model. The features with coefficients value of zero for across all the cluster-based models have been removed from the heatmap for better visual (33 zero-coefficient features)

**Supplementary Figure 5:**
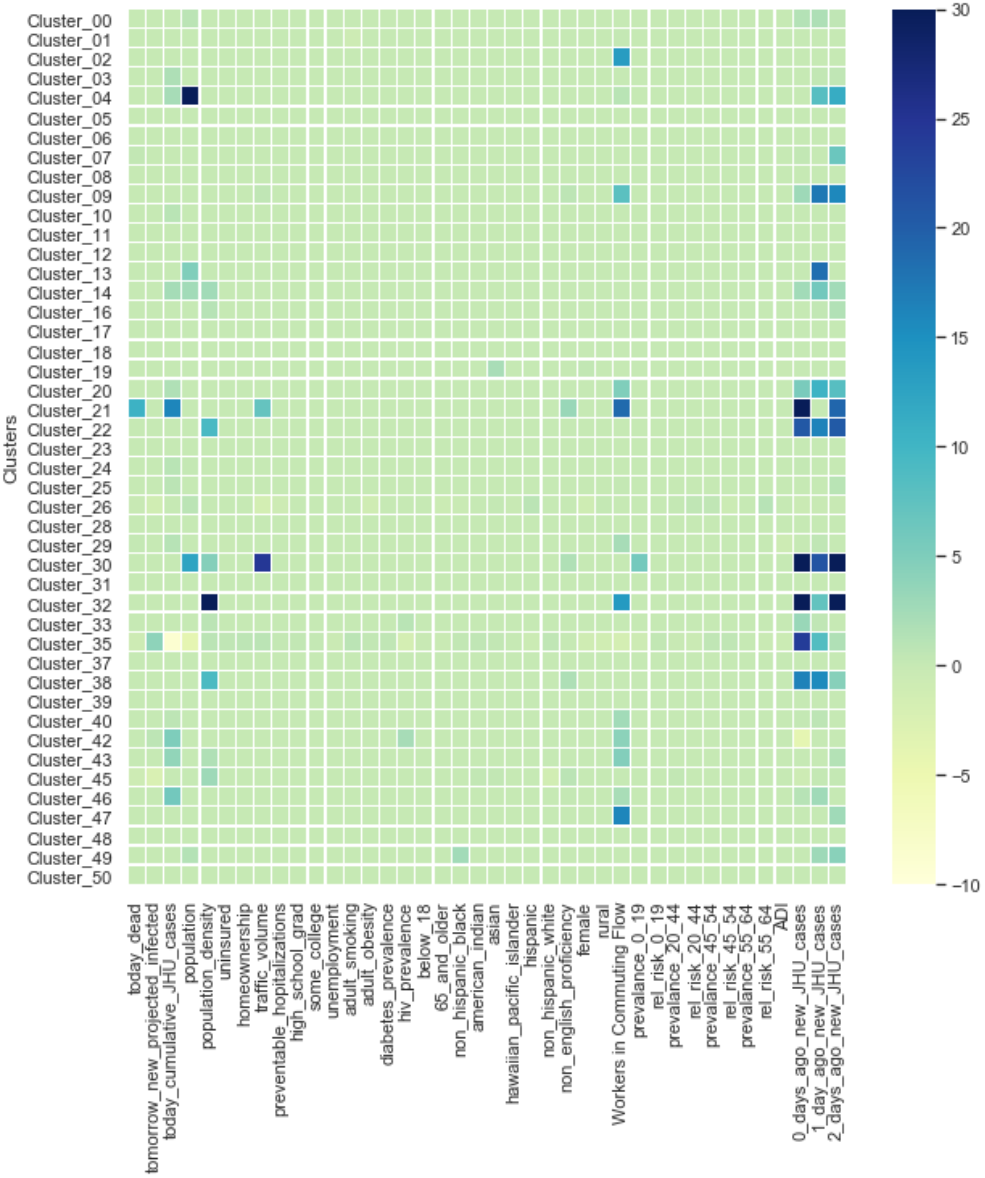
LASSO selected features for the last trained model (out of 14 date-dependents models) of each cluster trained for the counties clustered together based on their state boundary. New York County is part of cluster 32, yielding best preforming model.

## APPENDIX III: Data sources

| Feature | Source | Note |
| --- | --- | --- |
| dead count | JHU | Daily report |
| New projected all infected | ActNow output for March-April version | Date t+1 values were extracted from model ran on date t |
| Cumulative COVID cases | JHU | Daily report |
| New COVID cases for date t-0 | JHU | t-0 cumulative cases subtracted from day before |
| New COVID cases for date t-1 | JHU | t-1 cumulative cases subtracted from day before |
| New COVID cases for date t-2 | JHU | t-2 cumulative cases subtracted from day before |
| Area Deprivation Index | University of Wisconsin | Year 2018 |
| Workers in commuting flow | United States Census Bureau | 2011-2015 ACS commuting flows |
| - Prevalence of respiratory diseases per county broken down by age groups,<br>- Relative risk scores reflective of comorbidities | IBM® Advantage Suite® | 2018 |
| - Population,<br>- population density,<br>- uninsured,<br>- homeownership,<br>- traffic-volume,<br>- preventable hospitalization,<br>- high-school grad,<br>- some college,<br>- unemployment,<br>- adult smoking,<br>- adult obesity,<br>- diabetes prevalence,<br>- HIV prevalence,<br>- Age below 18,<br>- Age above 65,<br>- Non-Hispanic black,<br>- American Indian,<br>- Asian,<br>- Hawaiian pacific islander<br>- Hispanic,<br>- Non-Hispanic white,<br>- Non-English proficiency,<br>- Female,<br>- Rural | County Health Rankings and Roadmaps | - Data compiled from CDC and other public agency reports.<br>Year 2018 |

## Footnotes

1 As of April 12th, 2020, the ActNow model has gone through an update and “asymptomatic individuals” category has been added to the model. However, the projections used in this study are the outputs of the model prior to this change and thus do not include asymptomatic cases.

## Notes

### Competing Interest Statement

The authors have declared no competing interest.

### Funding Statement

No external funding was received.

### Author Declarations

The IRB exemption decision for this study was ruled by Western Institutional Review Board per below: "We determined this study is exempt from IRB review because it does not meet the definition of human subject research as defined in 45 CFR 46.102. Specifically, this project involves analysis of data from publicly available datasets and deidentified private datasets. The research activities do not involve human subjects, because the activities do not involve interaction or intervention with the subjects. Additionally, the investigator will not be able to readily ascertain the identity of any of the human subjects whose data is used in this project."

